# Early life factors and dementia risk: A study of adverse childhood experiences and later-life cognition and behaviour

**DOI:** 10.1101/2024.10.19.24315107

**Authors:** Dinithi Mudalige, Dylan X. Guan, Clive Ballard, Byron Creese, Anne Corbett, Ellie Pickering, Adam Hampshire, Pamela Roach, Eric E. Smith, Zahinoor Ismail

## Abstract

**Introduction:** Adverse childhood experiences (ACEs) are associated with brain alterations and cognitive decline. In later life, cognitive impairment and mild behavioural impairment (MBI) are associated with greater dementia risk.

**Objective & Study Design:** We investigated whether more severe ACEs are cross-sectionally associated with worse later-life cognitive and behavioural symptoms.

**Method:** Data are from the Canadian Platform for Research Online to Investigate Health, Quality of Life, Cognition, Behaviour, Function, and Caregiving in Aging (CAN-PROTECT). Measures included the Childhood Trauma Screener (CTS-5), neuropsychological testing, Everyday Cognition (ECog)-II scale, and MBI Checklist (MBI-C). Linear regressions modelled associations between ACEs severity and neuropsychological test scores. Multivariable negative binomial regressions (zero-inflated, if appropriate) modelled associations between ACEs severity and ECog-II and MBI-C scores. All models controlled for age, sex, education, and ethnocultural origin. Clinical diagnoses of depression and/or anxiety were explored as covariates or mediators.

**Results:** In adjusted analyses, higher ACEs scores were associated with worse performance on Trail-Making B (standardized b=0.10, q=0.003), Switching Stroop (b=-0.08, q=0.027), Paired Associates Learning (b=-0.08, q=0.049), and Digit Span (b=-0.08, q=0.029). Higher ACEs scores were also associated with higher ECog-II (b=1.08, q=0.029) and MBI-C (b=1.20, q<0.001) scores; these associations were neither mediated by affective symptoms (ECog p=0.16; MBI p=0.13) nor moderated by sex (ECog p=0.09; MBI p=0.46).

**Conclusion:** Older adults with a history of more severe ACEs show greater cognitive and behavioural risk markers for dementia that cannot be explained by previous psychiatric history. Further research into ACEs as an early modifiable risk factor for dementia is warranted.

## INTRODUCTION

Risk for dementia starts developing in early life ^1^, although beyond education, little emphasis has been placed on early-life factors such as adverse childhood experiences (ACEs). Of the 12 modifiable risk factors of dementia established by the Lancet Commission, only one – level of education – spans early life stages ^2^. However, early developmental stages involve extensive brain changes, and childhood experiences have the potential to have long-lasting effects. Because of this temporal association, targeting early-life factors may have a greater impact on dementia risk reduction than addressing modifiable risk factors in later life.

ACEs include abuse (physical, sexual, emotional), neglect, and household dysfunction occurring before the age of 18 ^3^. Studies have found long-term alterations in brain circuitry and structure secondary to ACEs. These alterations include amygdalar hyperactivity and volume reduction in the hippocampus, cerebellum, corpus callosum, and whole brain ^4–7^. These changes may lead to lower brain reserve, which is the brain’s physical capacity to maintain function despite deterioration. Those with lower brain reserve show clinical impairment with less neurological compromise (e.g., brain volume loss and/or neural dysconnectivity) compared to those with greater reserve ^8^. For example, an individual with ACEs-related hippocampal atrophy may experience earlier and more severe impairment from the changes in early-stage Alzheimer disease (AD) ^9^. Studies have found that those with ACEs have a greater risk of developing dementia ^10, 11^, indicating that the impact of ACEs can extend beyond the immediate aftermath of these experiences. ACEs can also be related to social, systemic, and structural determinants of health. For instance, the Lancet Commission reports that risk of cognitive decline and dementia are particularly high in socially disadvantaged populations ^2^.

However, beyond dichotomizing ACEs as present or absent, ACEs severity may be important to assess; few studies have taken this approach ^10, 12^. Specifically, ACEs severity may correspond to differential effects on brain function and varying degrees of dementia risk. Thus, incorporating ACEs severity and later-life dementia markers may provide insights into screening for those at risk of dementia.

In light of this, we examined two dementia markers: cognition and behaviour. Later-life changes in both objective and subjective measures of cognition are associated with a greater risk of incident dementia ^13, 14^. Similarly, later-life emergent and persistent changes in behaviour or personality are also associated with cognitive decline and incident dementia ^15^. Mild behavioural impairment is a syndrome that operationalizes later-life behavioural changes for dementia risk determination. MBI symptoms are categorized into five domains: decreased drive and motivation (apathy), affective dysregulation (mood and anxiety symptoms), impulse dyscontrol (agitation, impulsivity, abnormal reward salience), social inappropriateness (impaired social cognition), and abnormal perception or thought content (hallucinations and delusions, i.e., psychotic symptoms). Studies have indicated that MBI is associated with cognitive impairment cross-sectionally ^16, 17^ and incident cognitive decline and dementia longitudinally in cognitively normal (CN) older adults ^18, 19^, mixed CN and mild cognitive impairment (MCI) samples ^20–26^ and in MCI ^27, 28^.

Therefore, this study aimed to determine the relationships between ACEs and later-life changes in cognition and behaviour. We hypothesized that individuals with greater number and severity of ACEs would 1) have lower neuropsychological test scores; 2) experience more later-life cognitive symptoms; and 3) experience a greater burden of MBI, all of which are linked to dementia risk.

## METHOD

### Study Design

Data were obtained from the Canadian Platform for Research Online to Investigate Health, Quality of Life, Cognition, Behavior, Function, and Caregiving in Aging (CAN-PROTECT) ^1^. CAN-PROTECT is a nationwide online longitudinal observational cohort study of dementia-free community-dwelling Canadian residents aged ≥18 years. CAN-PROTECT consists of a validated neuropsychological test battery ^29^, detailed demographics, and participant-rated questionnaires on cognition, behaviour, function, quality of life, medical and mental health history, and lifestyle, amongst others. Currently, only baseline data are available. Extensive descriptions of CAN-PROTECT recruitment and data collection procedures have been published elsewhere ^1^. Participants provided informed consent as part of the registration process. Ethics approval for the study was acquired from the Conjoint Health Research Ethics Board at the University of Calgary.

### Participants

The overall sample from the CAN-PROTECT November 2023 data release comprised 2304 participants. Participants were excluded from the present analyses if they (1) had not yet completed the cognitive test battery (n=614); (2) did not complete questionnaires related to ACEs (n=306), subjective cognitive function (n=112), or MBI (n=47); or (3) or were younger than 50 years old (n=123) (Figure 1). Demographics between participants included and excluded for completing the cognitive battery, and those included and excluded for the completion of ACEs questionaries are indicated in Supplementary Figures 1 and 2, respectively. Only participants who were older than 50 years were included, in accordance with the MBI criterion stipulating the de novo onset of persistent neuropsychiatric symptoms (NPS) after the age of 50^15^. Therefore, the final sample size for analysis was 1102.

**Figure 1.**
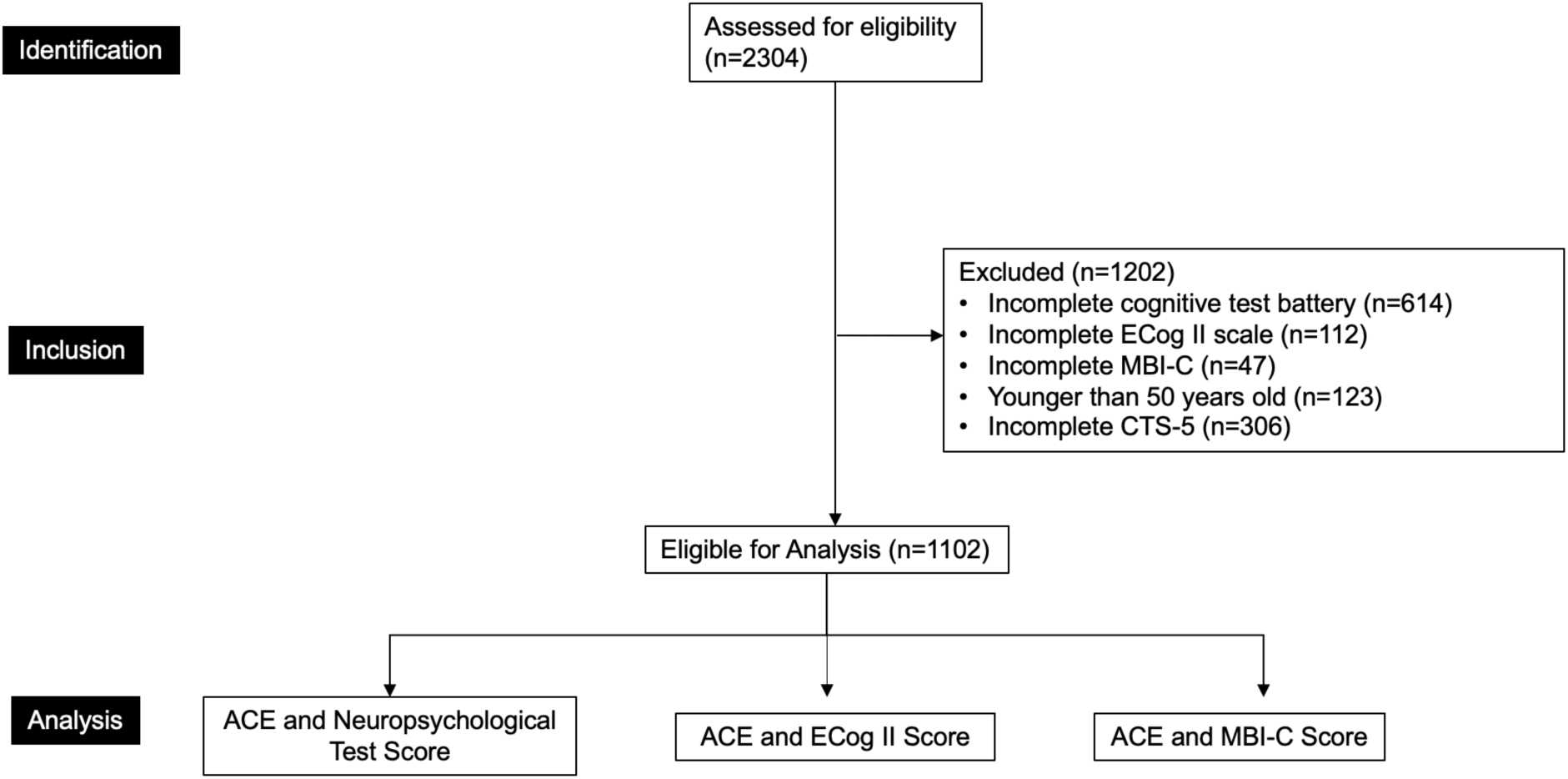
Participant Flow-Diagram. Neuropsychological tests include Trail-Making B, Switching Stroop, Self-Ordered Search, Paired Associate Learning, Digit Span, and Verbal Reasoning tasks. Abbreviations: CTS-5, Childhood Trauma Screener 5-Question Questionnaire; ACEs, Adverse Childhood Experiences; MBI-C, Mild Behavioral Impairment Checklist; ECog II, Everyday Cognition II.

### Measures

#### Adverse Childhood Experiences

ACEs were measured using the validated Childhood Trauma Screener 5-Question Questionnaire (CTS-5) ^30^. Participants responded with the frequency of the following on a scale ranging from 0 to 4, with 4 being very often true: “I felt loved”; “people from my family hit me so hard that I got bruises or scrapes”; “I felt like someone in my family hated me”; “someone sexually molested me”; and “there was someone to take me to the doctor when I needed it.” As the items “I felt loved” and “there was someone to take me to the doctor when I needed it” indicate more positive childhood experiences, these items were reverse-coded. Then, the responses to these five questions were summed to create the total ACEs severity score (range=0-20), where higher scores indicate more ACEs.

#### Neuropsychological Tests

Objective cognitive performance was measured using a validated neuropsychological test battery. This battery assesses performance on the Trail-Making B, Switching Stroop, Self-Ordered Search, Paired Associate Learning, Digit Span, and Verbal Reasoning tasks. All tasks, with the exception of Trail-Making B and Switching Stroop, are part of the Factors of Longitudinal Attention, Memory, and Executive Function cognitive battery ^29^ and have been developed and validated in online settings ^31^. Together, the battery measures cognitive domains of executive function, attention, task-switching, visual episodic memory, verbal reasoning, and working memory ^32–37^.

#### Everyday Cognition

Cognitive symptoms were measured with the self-reported Everyday Cognition (ECog)-II scale ^38^. The ECog-II asks participants to compare their cognition now to what it was 10 years ago with 41 questions across the domains of memory (9 questions), language (9 questions), visuospatial and perceptual abilities (8 questions), and executive functioning (15 questions).

Each question can be answered on a scale from 0-3, with 0 indicating no change and 3 indicating the most consistent change compared to 10 years ago. The ECog-II total score, ranging from 0-123, is calculated by summing the scores for each question, with higher scores indicating more severe symptoms.

#### Mild Behavioural Impairment

MBI was measured by the self-reported MBI-C, developed to operationalize the MBI-C criteria of later-life onset of persistent NPS that represent a change from longstanding behaviours^39^. The MBI-C comprises 34 items across five domains: decreased motivation (6 questions), emotional dysregulation (6 questions), impulse dyscontrol (12 questions), social inappropriateness (5 questions), and abnormal perception or thought content (5 questions). Each question assesses the presence and severity of symptoms, rated on a scale from 1 to 3 if present, with 3 indicating the greatest severity. The MBI total score, ranging from 0-102, is calculated by summing the scores for each question, with higher scores indicating greater global severity.

### Statistical Analysis

Participant demographics were summarized using descriptive statistics (means, standard deviations [SD], ranges, counts, and percentages). To compare groups, Wilcoxon signed-rank tests and chi-squared tests were used.

Multivariable linear regression modelled the association between ACEs (predictor) and neuropsychological test scores (outcome). Multivariable negative binomial regression modelled the association between ACEs (predictor) and ECog-II score (outcome), since the data were overdispersed (variance >> mean) counts. Multivariable zero-inflated negative binomial regression modelled the association between ACEs (predictor) and MBI-C score (outcome). All models were adjusted for age, sex, highest level of education, ethnocultural origin, and self-reported clinical diagnosis mood symptoms (depression and/or anxiety).

The associations between ACEs (predictor), ECog-II (outcome) score and MBI-C (outcome) were explored further through several secondary analyses. Two analyses explored to what extent affective symptoms mediated the associations between ACEs, ECog, and MBI-C, using bootstrapping with 1000 simulations. Moderation by sex was also examined for these associations. Lastly, the associations between ACEs (predictor) and MBI domains (outcomes) were explored using five multivariable negative binomial regressions. R version 4.0.5 was used to conduct all statistical analyses ^40^.

## RESULTS

### Participant Characteristics

Participant characteristics for all analyses, stratified by sex, have been summarized in Table 1. Of the total sample (n=1102), study participants were 64.6 (SD=7.3, range=50-89) years old on average, 883 (80.1%) were female, 290 (26.3%) had a bachelor’s degree or certificate equivalent to a bachelor’s degree, and 939 (85.2%) were of European origins. Additionally, 321 (29.1%) reported a previous diagnosis of depression, and 229 (20.8%) reported a previous diagnosis of anxiety. The average ACEs score was 2.05 (SD=2.7, range=0-19.0).

**Table 1.** Participant characteristics.

|  | **Total**<br>**(n=1102)** | **Male**<br>**(n=219)** | **Female**<br>**(n=883)** | ***p*** |
| --- | --- | --- | --- | --- |
| **Age (Years)** | 64.6 (7.3), 50.0-89.0 | 65.7 (7.8), 50.0-89.0 | 64.3 (7.2), 50.0-88.0 | 0.028<sup>c</sup> |
| **Education** |  |  |  | <0.001<sup>w</sup> |
| Some primary school | 1 (0.1) | 0 (0) | 1 (0.1) |  |
| Some high school | 12 (1.1) | 2 (0.9) | 10 (1.1) |  |
| High school diploma | 117 (10.6) | 25 (11.4) | 92 (10.4) |  |
| Trade certificate/diploma | 63 (5.7) | 27 (12.3) | 36 (4.1) |  |
| Non-university certificate/diploma | 276 (25.0) | 36 (16.4) | 240 (27.2) |  |
| University certificate below bachelor's | 55 (5.0) | 6 (2.7) | 49 (5.5) |  |
| Bachelor's degree | 290 (26.3) | 54 (24.7) | 236 (26.7) |  |
| degree/certificate above bachelor's | 288 (26.1) | 69 (31.5) | 219 (24.8) |  |
| **Ethno-Cultural Origin** |  |  |  | 0.33<sup>w</sup> |
| European Origins | 939 (85.2) | 182 (83.1) | 757 (85.7) |  |
| Non-European Origins | 163 (14.8) | 37 (16.9) | 126 (14.3) |  |
| **Mood Symptoms** |  |  |  | <0.001<sup>w</sup> |
| Absent | 696 (63.2%) | 161 (73.5%) | 535 (60.6%) |  |
| Present | 406 (36.8%) | 58 (26.5%) | 348 (39.4%) |  |
| **ACEs Score** | 2.05 (2.7), 0-19.0 | 1.6 (2.2), 0-12.0 | 2.2 (2.8), 0-19.0 | 0.012<sup>c</sup> |
| **ECog Score** | 11.7 (10.9), 0-88.0 | 11.6 (11.3), 0-63.0 | 11.7 (10.8), 0-88.0 | 0.71<sup>c</sup> |
| **Trail-Making B** | 60.9 (19.1), 26.8-193 | 63.2 (18.7), 34.4-148 | 60.4 (19.2), 26.8-193 | 0.016<sup>c</sup> |
| **Switching Stroop** | 38.5 (14.3), 0-83.3 | 39.6 (14.4), 3.00-75.0 | 38.3 (14.3), 0-83.3 | 0.19<sup>c</sup> |
| **Self-Ordered Search** | 6.7 (2.5), 0-14.0 | 7.0 (2.7), 0-12.0 | 6.6 (2.5), 0-14.0 | <0.001<sup>c</sup> |
| **Paired Associates Learning** | 3.9 (0.9), 0-7.0 | 3.8 (0.9), 0-7.0 | 4.0 (0.9), 0-7.0 | 0.019<sup>c</sup> |
| **Verbal Reasoning** | 32.0 (9.4), -1.0-67.0 | 31.1 (9.2), -1.0-61.3 | 32.2 (9.5), 0-67.0 | 0.060<sup>c</sup> |
| **Digit Span** | 7.0 (1.8), 0-20.0 | 6.9 (1.7), 0-16.0 | 7.0 (1.8), 0-20.0 | 0.51<sup>c</sup> |
| **MBI Score** | 5.3 (7.4), 0-65.0 | 4.9 (7.0), 0-49.0 | 5.4 (7.5), 0-65.0 | 0.25<sup>c</sup> |

*Note.* All values for continuous variables are shown in mean (SD), range. Values were rounded to one decimal place where appropriate. Categorical variables are shown as n (%). Participants missing data in any of the variables were excluded from descriptive statistical analysis.
Abbreviations: ECog-II, everyday cognition; MBI, mild behavioral impairment; ACEs, adverse childhood experience w – Wilcoxon Rank Sum (Mann-Whitney) Test; c – Chi-Squared Test

### Adverse Childhood Experiences and Cognition

For objective neuropsychological test scores, every 1 SD increase in ACEs score was associated with an increase of 0.10 SD (95%CI:0.05 – 0.16, q=0.003) on Trail-Making B score, and decreases of 0.08 SD (95%CI:-0.14– -0.03, q=0.027) on Switching Stroop, Paired Associates Learning (95%CI:-0.13 – -0.02, q=0.049), and Digit Span (95%CI:-0.14 – -0.02, q=0.029) scores. Self-Ordered Search (b=-0.05, 95%CI:-0.11– -0.01, q=0.22) and Verbal Reasoning Tasks (b=-0.05, 95%CI:-0.11– -0.01, q=0.22) did not demonstrate statistically significant associations with ACEs score, although the magnitude and direction of effect were similar to the other neuropsychological tests.

For subjective cognition, every 1 SD increase in ACEs score was associated with a 1.08-fold (95%CI:1.02 – 1.14, q=0.029) higher ECog-II score, adjusting for all covariates (Table 2). This association was neither mediated by mood symptoms (b=0, 95%CI:0 – 0.01, p=0.16) nor moderated by sex (b=0.14, 95%CI:-0.04 – 0.31, p=0.09). Scatterplot distributions for these associations are illustrated in Supplementary Figures 1 and 2.

**Table 2.**
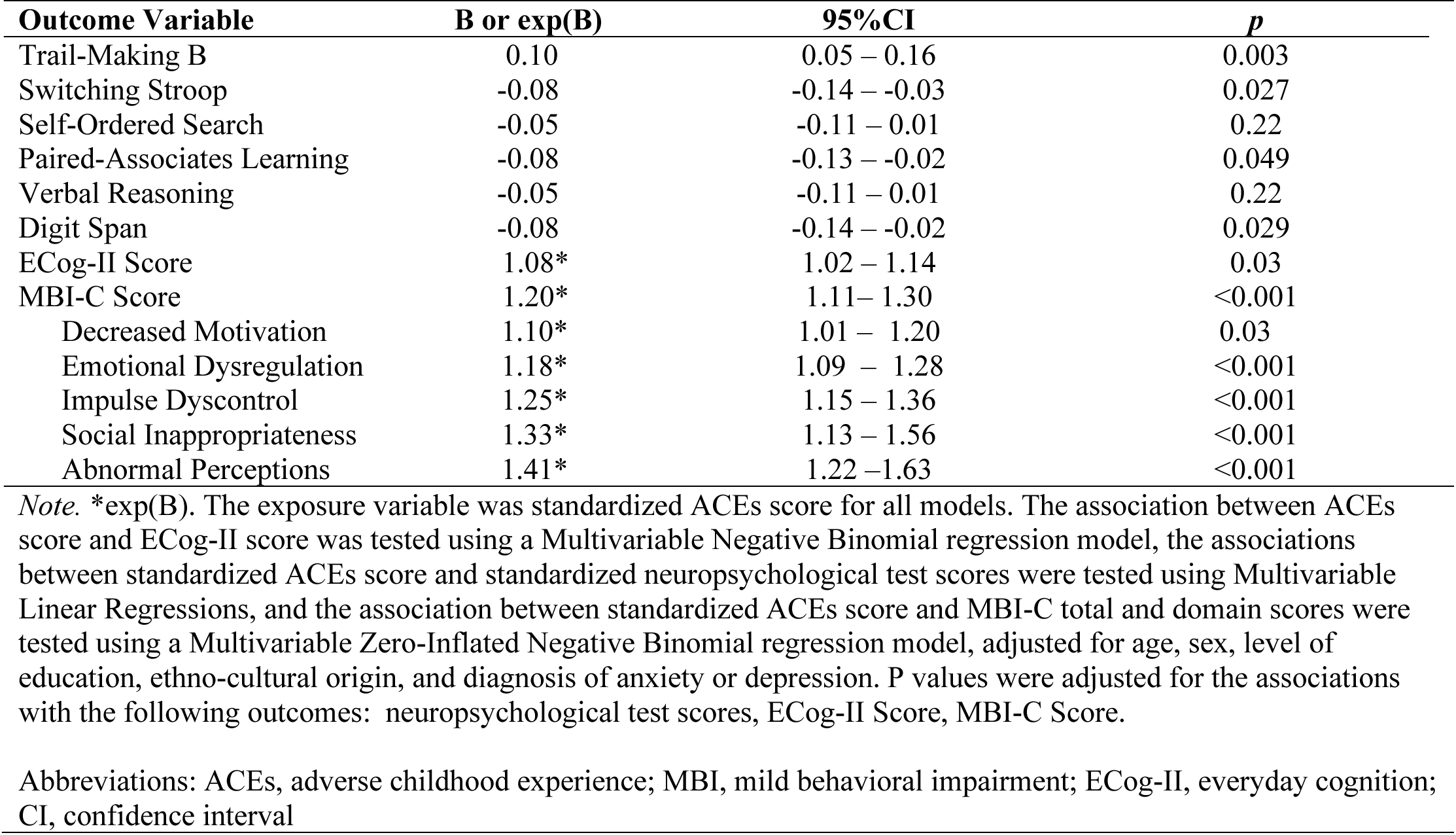
Multivariable regression models for the association between ACEs score and ECog-II, neuropsychological tests scores, and MBI-C scores.

### Adverse Childhood Experiences and Mild Behavioural Impairment

For total MBI, every 1 SD increase in the ACEs score was associated with a 1.20-fold (95%CI:1.11 – 1.30, q<0.001) higher MBI-C score, adjusting for all covariates (Table 2).

Additionally, this association was neither mediated by mood symptoms (b=0.12, 95%CI:-0.04 – 0.29, p=0.13) nor moderated by sex (b=0.11, 95%CI:-0.11 – 0.32, p=0.46). A scatterplot distribution for this association is illustrated in Supplementary Figure 3.

For MBI domains, every 1 SD increase in ACEs score was associated with a 10.2% (95%CI: 1.1 – 20.2, p= 0.027) higher decreased motivation, 18.4% (95%CI: 9.3 – 28.2, p<0.001) higher emotional dysregulation, 25.1% (95%CI: 14.7 – 36.4, p<0.001) higher impulse dyscontrol, 33.2% (95%CI: 13.6 – 56.3, p<0.001) higher social inappropriateness, and 40.7% (95%CI: 21.6 – 62.8, p<0.001) higher abnormal perceptions or thought content score.

## DISCUSSION

This study explored associations between experiences in childhood and risk for dementia in later life. As hypothesized, more severe ACEs were found to be associated with more severe cognitive and behavioural symptoms, both of which reflect a greater risk for incident dementia. These symptoms could not be explained by previous psychiatric history. Our findings emphasize the role of quantifying the severity of ACEs in determining dementia risk using cognition and behaviour. This study adds depth to our understanding of childhood experiences as a component of social, systemic, and structural determinants of later-life brain health.

The fact that ACEs are associated with worse cognition is consistent with the current literature. For instance, in community-dwelling older adults, those with a history of ACEs were more likely to experience cognitive impairment ^41, 42^. However, the links to dementia are less clear. In a French cohort, while ACEs were associated with psychomotor slowing, especially in females, there was no association with incident dementia ^43^. In a sample of former male professional football players with a mean age of 57.2 (SD=13.5, range=28-92), ACEs were associated with cognitive and functional impairment consistent with dementia based on the AD8 scale ^10^. Similarly, another study in Japanese older adults found that those who experienced three or more ACEs had a greater risk of developing dementia than those who grew up without ACEs, controlling for age, sex, childhood economic hardship, nutritional environment, and education ^11^.

Exploring the link between ACEs and NPS is more novel and studied less often, especially in the context of dementia risk. In the same study of football players, ACEs were associated with probable depression (measured using the PHQ-9). However, after adjusting for concussions, depression no longer differed from those without ACEs ^10^. Another study found that in Chinese older adults, those with ACEs were more likely to have psychiatric disorders in the past year after adjusting for age, sex, marital status, employment status, education, rural or urban residence, region, and physical diseases ^44^. However, to our knowledge, there are no data on the relationship between ACEs and MBI, which is a novel but well-validated dementia risk syndrome.

Nonetheless, ACEs and MBI both indicate greater risk for incident dementia^11, 15^. It is plausible that ACEs contribute to greater psychiatric symptoms, which then contribute to cognitive impairment and dementia risk. However, our finding that neither the association between ACEs and cognition nor the association between ACEs and MBI were mediated by previous psychiatric history indicates that these relationships may involve other pathways unrelated to psychiatric conditions.

An alternative hypothesis may involve brain reserve. Subtypes of ACEs, including childhood abuse and neglect, have been linked to inhibited blood flow to the prefrontal cortex, impeding development ^7^. These changes have subsequently been linked to poorer executive functioning ^45^. As the prefrontal cortex is a critical region associated with many NPS ^46^, reduced prefrontal cortex volume and thickness (indicating reduced brain reserve), may also be associated with more severe NPS. Moreover, childhood abuse is also associated with elevated cortisol ^47^.

Cortisol has been found to promote amyloid β plaque deposits ^48, 49^, one of the hallmark features of AD. The accumulation of these plaques is related to brain atrophy, linked to worsening cognitive and behavioural symptoms as individuals age ^50, 51^. In fact, a recent study has found that while total stressful life events were not associated with AD pathophysiology and neuroinflammation, stressful life events occurring in childhood were ^52^. Additionally, ACEs may be related to heightened levels of systemic inflammation, which presents another potential mechanism for the association between ACEs-related cognitive and behavioural impairment.

Studies have found associations between higher C-reactive protein (CRP) levels, a marker of systemic inflammation ^53^, and poorer performance on tests of memory ^54, 55^. Similarly, inflammation plays an important role in the pathogenesis of neuropsychiatric disorders ^56^.

Neuroinflammation may be implicated in neurobehavioural changes, including the development of neuropsychiatric disorders ^57^. ACEs are also linked with lower academic achievement and pursuit of higher education ^58, 59^. As education contributes to cognitive reserve and is therefore protective against cognitive decline ^60^, those with more severe ACEs may be less protected against cognitive and behavioural impairment and onset of dementia.

These findings emphasize that those with ACEs are at heightened risk for dementia, although further longitudinal data are required to understand risk in this specific sample. Nonetheless, clinicians might consider a dementia risk assessment, including measures of later-life changes in cognition and behaviour, in older adults who present with a history of ACEs. If addressing ACEs can prevent or mitigate later-life cognitive and behavioural impairment and decrease the risk of dementia, then ACEs might be considered a modifiable risk factor. At a public health level, measures to mitigate ACEs could be amongst the earliest of dementia prevention interventions. In addition to considering childhood public health and policy through a dementia risk factor lens, these findings prompt further investigations to determine the mechanisms involved in the association between ACEs and cognitive and behavioural risk markers for dementia. Of note, our sample comprises mostly cognitively unimpaired participants, none with dementia diagnoses or living in continuing care settings.

There are limitations to our study. These are cross-sectional data, so no causal associations can be inferred. Further, the five-item questionnaire that assessed ACEs may not encompass all domains of childhood adversity. For instance, domains of ACEs, such as witnessing interpersonal violence, family substance use, family incarceration, or family mental illness, are not adequately captured by the five questions in the CTS-5. Additionally, the ECog-II is a subjective measure of cognition and may not accurately depict objective cognitive status. In light of this, we used objective neuropsychological tests to assess domains of cognition and differences in performance were found, which were consistent with the subjective cognitive reports. Lastly, other mediating factors aside from previous psychiatric history may have contributed to the relationship between ACEs and cognition/MBI. Future studies could assess additional aspects of ACEs to examine whether other factors contribute to the aforementioned relationships and explore the underlying determinants of ACEs, cognition, and MBI.

Regardless, we establish a starting point for further investigation regarding this novel relationship between ACEs severity and later-life cognitive and behavioural impairment. Additionally, this study emphasizes the importance of early-life factors for the risk of dementia and is relevant at a public health level. Further, the large sample size also provided sufficient statistical power to test several hypotheses, including moderation and mediation, enabling a fuller understanding of the relationship between ACEs, cognition, and MBI. As longitudinal data from CAN-PROTECT become available, we will be able to assess change over time and build on these foundational results.

## CONCLUSION

This study demonstrated that older adults with a history of ACEs have more cognitive and behavioural symptoms in later life, indicative of greater dementia risk. These relationships were not mediated by previous psychiatric history, indicating that ACEs may directly contribute to later-life cognitive and behavioural impairment. The relevance of our study is reflected by the novelty of the findings, the identification of an at-risk sub-population of older adults who can be screened for dementia risk with assessments of cognitive and behavioural change, and the potential to inform preventative strategies aimed at reducing the burden of dementia, starting in childhood.

## Data Availability

All data produced in the present study are available upon reasonable request to the authors.

## SUPPLEMENTARY TABLES/FIGURES

**Supplementary Table 1.** Participant Characteristics for Incomplete and Complete Cognitive Battery.

|  | **Total**<br>**(N=2304)** | **Cognitive Battery**<br>**Incomplete**<br>**(N=614)** | **Cognitive Battery**<br>**Complete**<br>**(N=1690)** | ***p*** |
| --- | --- | --- | --- | --- |
| **Age** | 62.8 (9.43), 23.0-91.0 | 62.3 (10.4), 34.0-88.0 | 63.0 (9.05), 23.0-91.0 | 0.22 |
| **Sex** |  |  |  | 0.55 |
| Male | 513 (22.3%) | 142 (23.1%) | 371 (22.0%) |  |
| Female | 1791 (77.7%) | 472 (76.9%) | 1319 (78.0%) |  |
| **Education** |  |  |  | 0.82 |
| Some primary school | 1 (0.0%) | 0 (0%) | 1 (0.1%) |  |
| Some high school | 32 (1.4%) | 8 (1.3%) | 24 (1.4%) |  |
| High school diploma | 246 (10.7%) | 72 (11.7%) | 174 (10.3%) |  |
| Trade certificate/diploma | 134 (5.8%) | 32 (5.2%) | 102 (6.0%) |  |
| Non-university certificate/diploma | 560 (24.3%) | 146 (23.8%) | 414 (24.5%) |  |
| University certificate below bachelor's | 131 (5.7%) | 41 (6.7%) | 90 (5.3%) |  |
| Bachelor's degree | 600 (26.0%) | 161 (26.2%) | 439 (26.0%) |  |
| degree/certificate above bachelor's | 600 (26.0%) | 154 (25.1%) | 446 (26.4%) |  |
| **Ethno-Cultural Origin** |  |  |  | 0.43 |
| European Origins | 1926 (83.6%) | 507 (82.6%) | 1419 (84.0%) |  |
| Non-European Origins | 378 (16.4%) | 107 (17.4%) | 271 (16.0%) |  |

*Note.* All values for continuous variables are shown in mean (SD), range. Values were rounded to one decimal place where appropriate.
w – Wilcoxon Rank Sum (Mann-Whitney) Test; c – Chi-Squared Test

**Supplementary Table 2.** Participant Characteristics for Participants with Childhood Trauma Screener Data Absent and Present.

|  | **Total**<br>**(N=1408)** | **CTS Data Absent**<br>**(N=306)** | **CTS Data Present**<br>**(N=1102)** | ***p*** |
| --- | --- | --- | --- | --- |
| **Age** | 64.5 (7.41), 50.0-89.0 | 64.1 (7.77), 50.0-88.0 | 64.6 (7.31), 50.0-89.0 | 0.20 |
| **Sex** |  |  |  | 0.37 |
| Male | 287 (20.4%) | 68 (22.2%) | 219 (19.9%) |  |
| Female | 1121 (79.6%) | 238 (77.8%) | 883 (80.1%) |  |
| **Education** |  |  |  | 0.42 |
| Some primary school | 1 (0.1%) | 0 (0%) | 1 (0.1%) |  |
| Some high school | 18 (1.3%) | 6 (2.0%) | 12 (1.1%) |  |
| High school diploma | 148 (10.5%) | 31 (10.1%) | 117 (10.6%) |  |
| Trade certificate/diploma | 76 (5.4%) | 13 (4.2%) | 63 (5.7%) |  |
| Non-university certificate/diploma | 365 (25.9%) | 89 (29.1%) | 276 (25.0%) |  |
| University certificate below bachelor's | 75 (5.3%) | 20 (6.5%) | 55 (5.0%) |  |
| Bachelor's degree | 357 (25.4%) | 67 (21.9%) | 290 (26.3%) |  |
| degree/certificate above bachelor's | 368 (26.1%) | 80 (26.1%) | 288 (26.1%) |  |
| **Ethno-Cultural Origin** |  |  |  | 0.08 |
| European Origins | 1187 (84.3%) | 248 (81.0%) | 939 (85.2%) |  |
| Non-European Origins | 221 (15.7%) | 58 (19.0%) | 163 (14.8%) |  |

*Note.* All values for continuous variables are shown in mean (SD), range. Values were rounded to one decimal place where appropriate.
Abbreviations: CTS; childhood trauma screener w – Wilcoxon Rank Sum (Mann-Whitney) Test; c – Chi-Squared Test

**Supplementary Figure 1.**
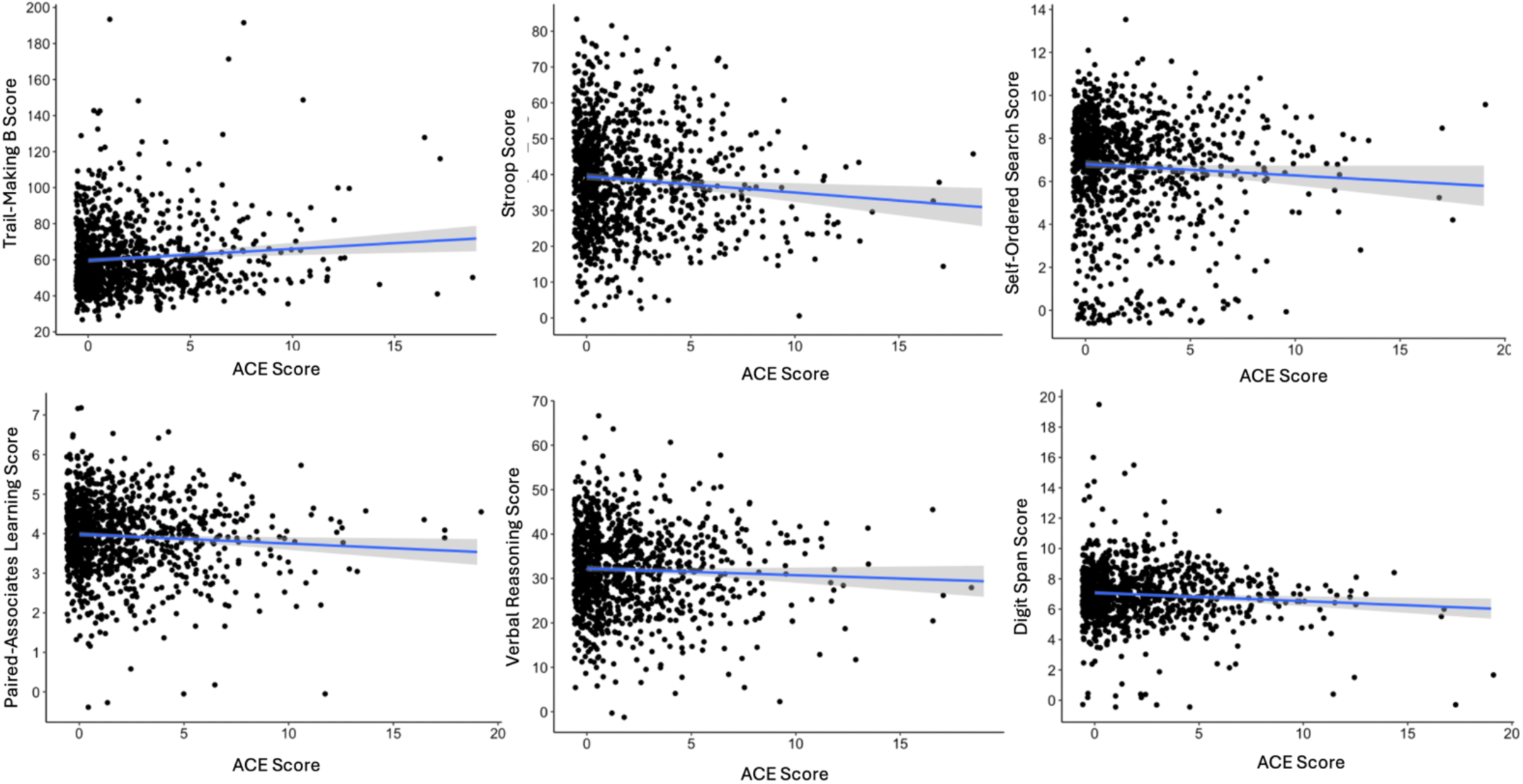
Scatterplot distributions. Adverse childhood experiences (ACEs) score as a function of neuropsychological test scores

**Supplementary Figure 2.**
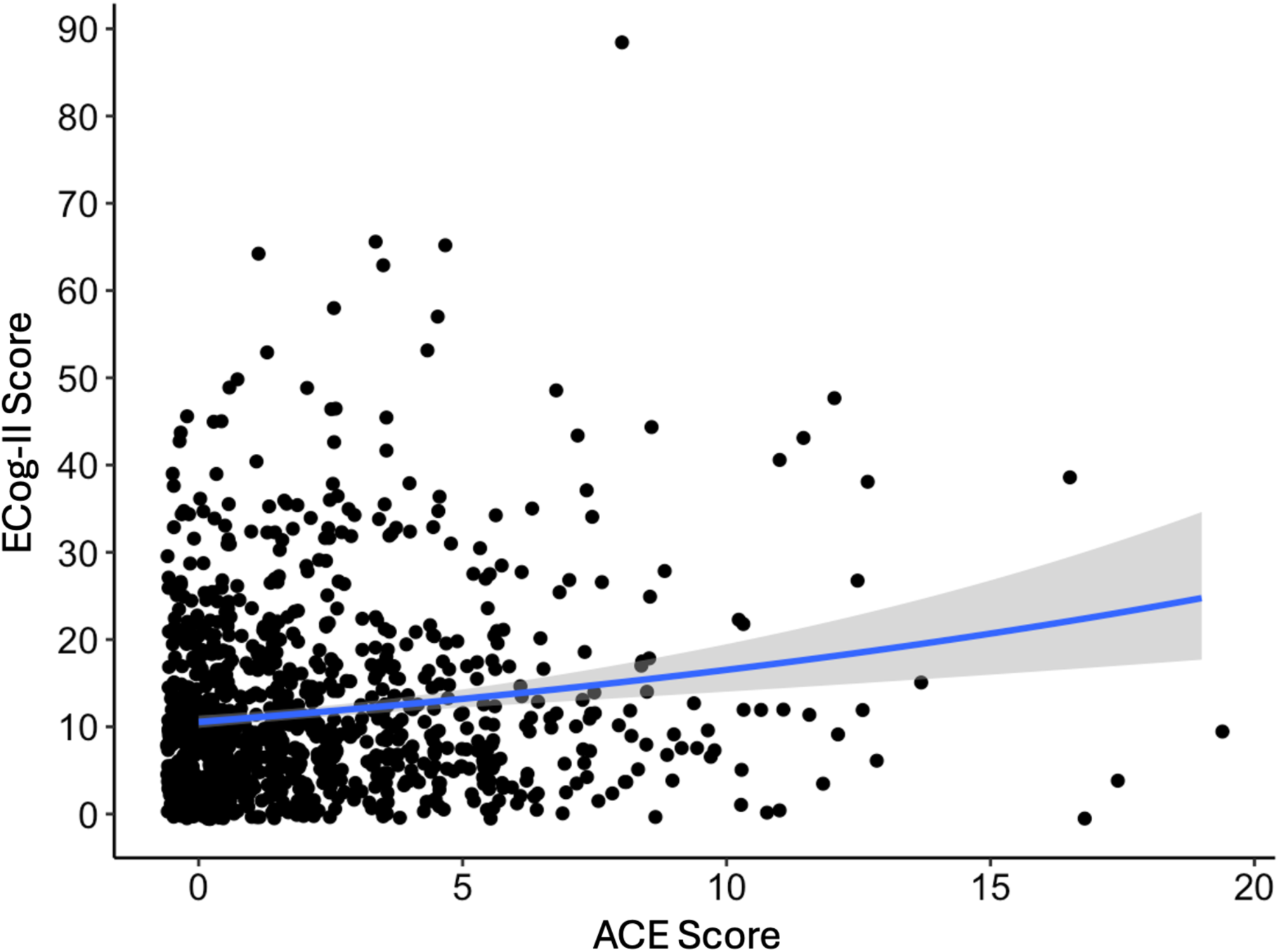
Scatterplot distribution. Adverse childhood experiences (ACEs) score as a function of everyday cognition II (ECog-II) score

**Supplementary Figure 3.**
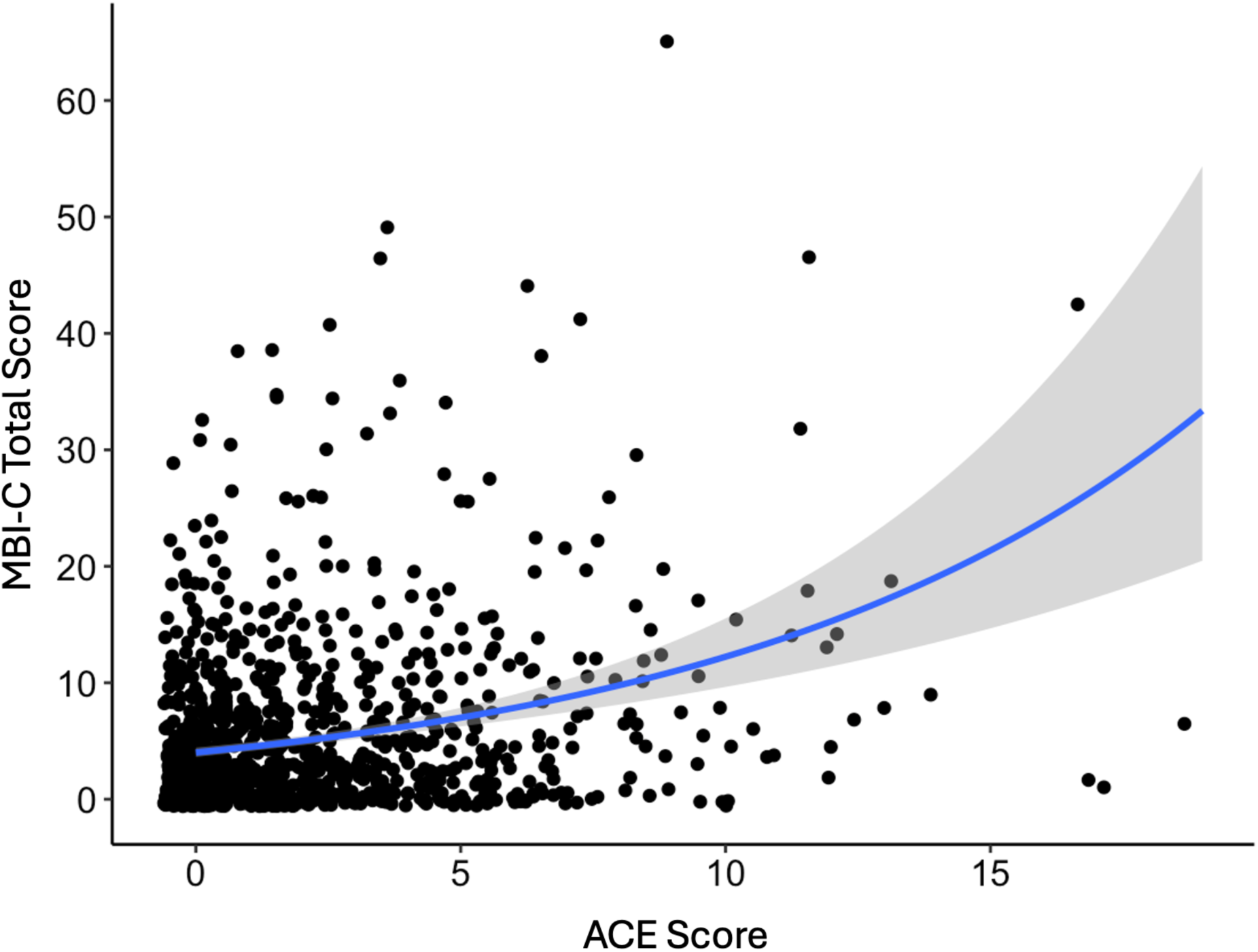
Scatterplot distribution. Adverse childhood experiences (ACEs) score as a function of mild behavioral impairment checklist (MBI-C) total score

